# Interventions with dietary supplements, including pre-, pro- and synbiotics, to reduce acute and late gastrointestinal side effects in patients undergoing pelvic radiotherapy: A systematic review and meta-analysis

**DOI:** 10.1101/2020.08.21.20178814

**Authors:** Benjamin Bartsch, Chee Kin Then, Elinor Harriss, Christiana Kartsonaki, Anne E. Kiltie

## Abstract

Pelvic radiotherapy (RT) often results in toxicity to the gastrointestinal tract and clinical trials have demonstrated a potential benefit of dietary supplements in alleviating acute effects. However, no prophylactic agents have been approved to date for relief of gastrointestinal side-effects caused by pelvic radiation. This systematic review and meta-analysis were undertaken with the aim of evaluating the efficacy of a number of dietary supplement interventions in preventing or alleviating symptoms of gastrointestinal toxicity in patients undergoing RT for a range of common pelvic malignancies. The search protocol was prospectively submitted to PROSPERO at the University of York. CENTRAL, MEDLINE, EMBASE, and ClinicalTrials.gov were searched up to June 2020 for randomised controlled clinical trials. Interventions included four supplement categories: biotics, amino acids, poly-unsaturated fatty acids and polyphenols. Efficacy was determined with reference to outcomes based on symptoms of acute gastrointestinal toxicity, including diarrhoea, nausea, vomiting, flatulence/bloating, bowel movement frequency, tenesmus and rectal bleeding. Twenty-three randomised controlled trials (1919 patients) were identified in this review. Compared with placebo, probiotics (RR=0.71; 95% CI: 0.52 to 0.99), synbiotics (RR=0.45; 95% CI: 0.28 to 0.73) and polyphenols (RR=0.30; 95% CI: 0.13 to 0.70) were significantly associated with a lower risk of diarrhoea. Biotic supplements also reduced the risk of moderate to severe diarrhoea (RR=0.49; 95% CI: 0.36 to 0.67) and the need for anti-diarrhoeal medication (RR=0.64; 95%CI: 0.44 to 0.92). In contrast, amino acid supplements had no effect on acute symptoms (RR=1.05; 95% CI:0.86 to 1.29). There was a non-significant trend for reduction in nausea and mean bowel movements per day using dietary supplements. Biotic supplements, especially probiotics and synbiotics, reduce acute symptoms of gastrointestinal toxicity in patients undergoing pelvic radiotherapy.

## INTRODUCTION

Radiotherapy is a major cancer treatment modality, used to treat approximately 50% of patients[1]. Chemoradiation (concurrent delivery of systemic chemotherapy) is generally preferred over radiotherapy alone for most pelvic malignancies, including tumours of the lower gastrointestinal, gynaecological and urological (with the exception of prostate cancer) tracts[2]. Over 200,000 patients in the US are treated with pelvic or abdominal radiotherapy each year[3]. It is inevitable that normal gastrointestinal tissues are exposed to radiation during pelvic radiotherapy[4], with approximately 80% of patients developing acute symptoms of radiation-induced gastrointestinal toxicity[5]. However, despite their impact on patients’ quality of life, no prophylactic agents for the alleviation of gastrointestinal side-effects from pelvic radiation have been approved to date[6].

Acute symptoms usually develop during or immediately after RT, and typically improve within three months following RT[7]. The most common acute side effect is diarrhoea, affecting up to 80% of all patients[8]. Other symptoms, such as abnormal stool output, vomiting, rectal bleeding, tenesmus and gastrointestinal discomfort are also common. Late symptoms include GI bleeding, fistula, stricture and colostomy[9].

Use of a dietary supplement is aimed at boosting daily intake of specific nutrients, to much higher levels than obtained from the diet, to alleviate symptoms of gastrointestinal toxicity. Such dietary supplements include biotics, amino acids, poly-unsaturated fatty acids (PUFAs) and polyphenols. Probiotics, mainly of the Lactobacillus and Bifidobacteria genera, are live microorganisms thought to produce health benefits following passage to the intestine[10]. Prebiotics are soluble or non-soluble dietary fibres, that pass undigested through the upper gastrointestinal tract and are metabolised by bacteria in the colon, thus enhancing gut microbiota beneficial to the host’s health[11]. The use of synbiotics refers to administration of a combination of prebiotics and probiotics; the presence of the prebiotic enhances survival of the probiotics in the lower gastrointestinal tract. Amino acids, poly-unsaturated acids (PUFAs) and polyphenolic compounds have also been employed in supplement intervention strategies in pelvic RT. Anti-inflammatory effects of the omega-6 PUFA conjugated linolenic acids are seen in inflammatory bowel disease[12]. Glutamine is the most abundant amino acid with important roles in support of mucosal growth and function. Evidence from animal studies confirms that dietary glutamine supplementation can protect the small intestinal epithelium from radiation damage[13]. Polyphenolic compounds extracted from plants, such as flavanols, flavandiols, epigallocatchin and genistein, can also suppress or alleviate radiation-induced intestinal injury.

This review tests the hypothesis that administration of oral dietary supplements for cancer patients receiving pelvic radiotherapy may trigger changes in the lower gastrointestinal tract which lead to a reduction in gastrointestinal toxicity. We conducted a systematic review and meta-analysis of randomised controlled trials (RCTs), with the aim of determining whether dietary interventions using supplements can alleviate symptoms of gastrointestinal toxicity in pelvic RT patients, including diarrhoea, nausea, vomiting, flatulence/bloating, bowel movement frequency, tenesmus and blood in bowel movements.

## METHODS AND MATERIALS

### Trial registration number

The study protocol was published on the PROSPERO international prospective register of systematic reviews (registration number CRD42020183304).

### Search strategy and study selection

The following electronic databases were searched from inception to the search date (19/06/2020) for relevant literature: Cochrane CENTRAL, Ovid Medline, Ovid Embase, and ClinicalTrials.gov. The search strategies included both medical subject heading and free text terms to retrieve relevant RCTs and non-randomised studies about the gastrointestinal side effects in cancer patients undergoing pelvic radiotherapy, limiting to studies in humans only. The full set of search strategies is available in Appendix 1 to 3, and inclusion and exclusion criteria are available in the PROSPERO registration[14]. Relevant articles were identified on PubMed. Handsearching of meta-analyses, systematic reviews and papers identified studies not indexed in the electronic databases used for this review. All titles and abstracts retrieved by electronic searches were downloaded and duplications removed using EndNote reference management software.

### Data extraction

Systematic data collection from included studies was conducted using a data collection form designed specifically for this review. It included the following information (where available) for each dataset: publication year, study design, participants (number, age distribution, gender distribution, details of malignancy, details relevant to inclusion and exclusion criteria), current cancer treatment, intervention and measured outcomes (diarrhoea incidence and severity and details of other GI symptoms recorded as toxicity scores, patient-reported outcome measure scores, questionnaires or interview scores).

### Outcome assessment

Different measures of treatment effects were used for dichotomous and continuous outcomes, namely, risk ratio (RR) for dichotomous outcomes and the mean difference between the intervention and control arms for continuous outcomes. Standardised mean difference was used to compare to compare results from studies that reported the same outcomes measured on different scales.

### Study quality, assessment of heterogeneity and publication bias

Risk of bias assessment was carried out for all studies that met the inclusion criteria, using the Cochrane Risk of Bias 2 tool. To assess the heterogeneity, we used a chi-squared test and I^2^. P values less than 0.1 were considered as evidence of heterogeneity. Tau-squared is the estimated standard deviation of underlying effects across studies. Begg’s funnel plots were used to visually assess asymmetry potentially due to publication bias.

### Data synthesis and statistical analysis

Meta-analyses were performed to measure the effect of dietary supplements on an outcome, in instances where there were three or more studies that reported the same outcome. All analyses were conducted using RevMan 5.4 and R version 4.0.2 with package ‘meta’. For dichotomous outcomes, RR were estimated and were meta-analysed using a random effects model using the Mantel-Haenszel method. For continuous outcomes, mean differences were estimated and were pooled using a random effects model with the inverse variance method. 95% confidence intervals (CI) for all estimates were calculated. Meta-regression by mean age, proportion of male participants and sample size was used to assess whether the effects of interventions on incidence of diarrhoea varied by these study characteristics.

## RESULTS

The search of the four primary databases identified 23,542 titles published between 1946 and June 2020 (search process summarised in Figure 1). After 5,825 duplications were removed, a total of 17,717 entries remained. These studies were manually reviewed by title and abstract and 17 met the inclusion criteria. Six further studies were identified from manual searches of the reference sections of research articles. Finally, 23 studies met the inclusion criteria and could be used for quantitative analysis. The effects of interventions on incidence of diarrhoea did not vary by mean age (p=0.552), proportion of male participants (p=0.131), sample size (p=0.131) or RT dose (p=0.073) (Figure S1). Results of the overall and individual risk of bias assessments for each of the five domains are presented in Figure 2.

**Figure 1.**
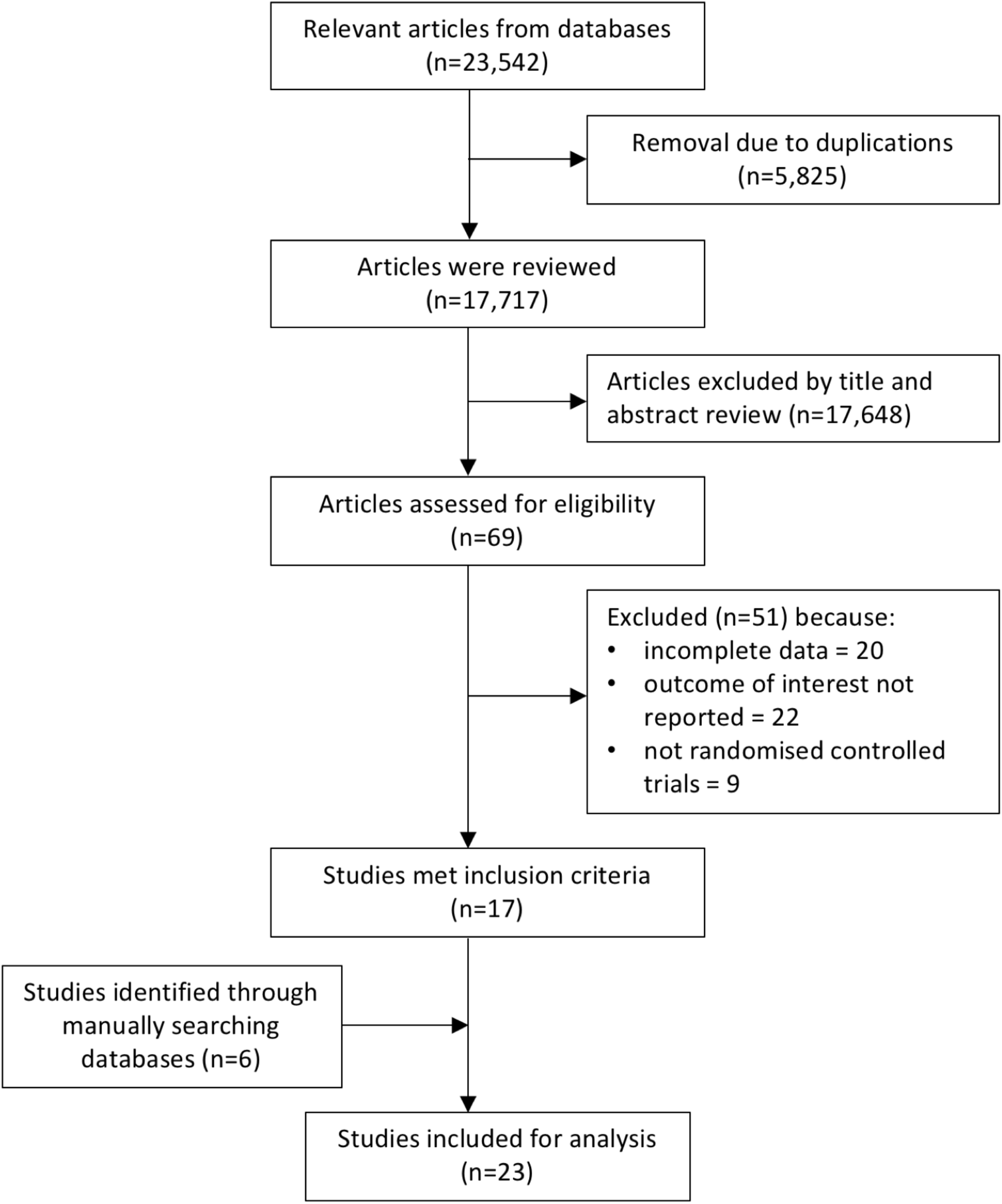
PRISMA flow chart of studies evaluated in the systematic review.

**Figure 2.**
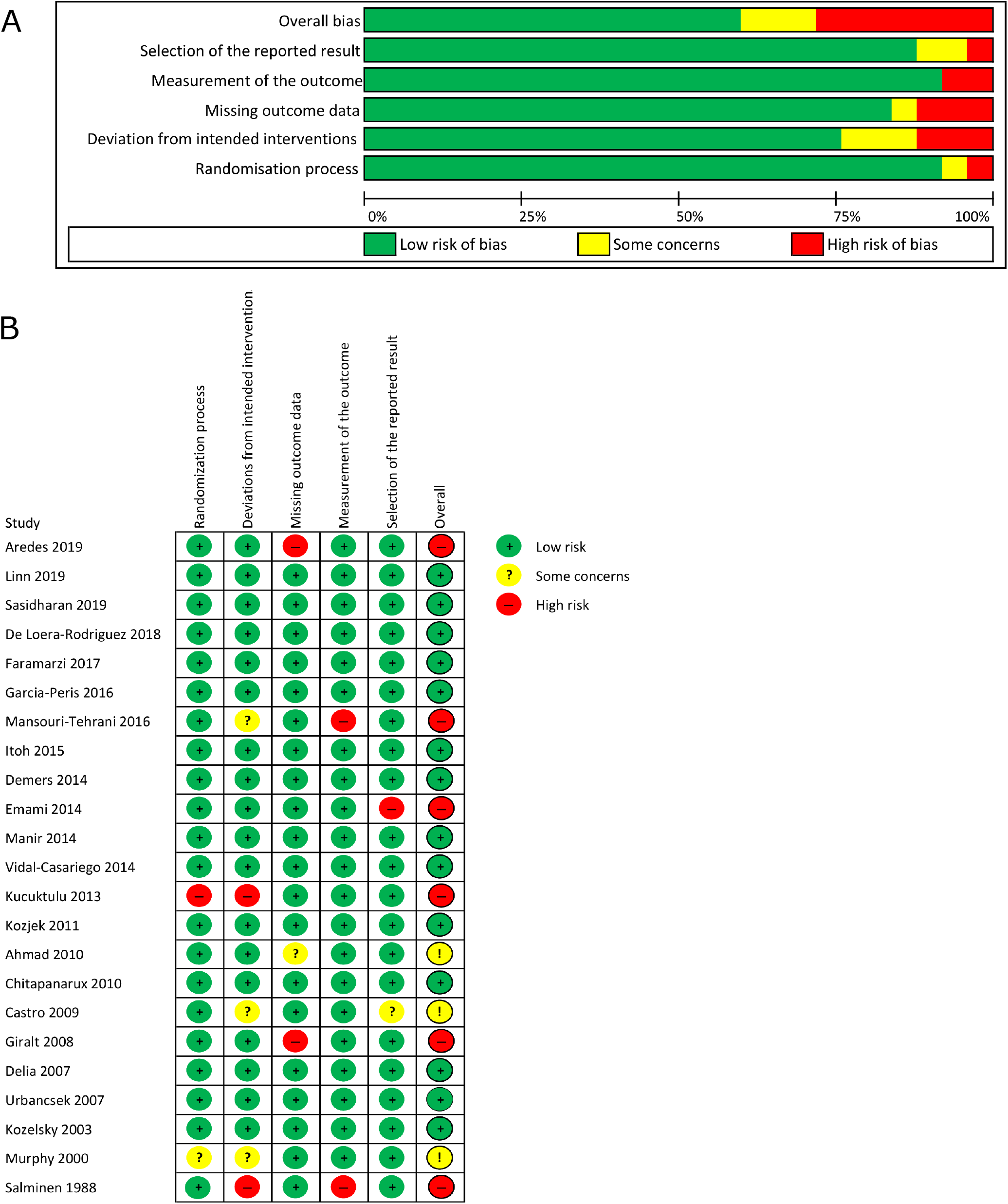
Risk of bias summary for all studies that met the inclusion criteria.

### Included studies and characteristics of included studies

In total 23 studies involving a total of 1,919 patients met the inclusion criteria and for each outcome, they were grouped by intervention category. These studies were all randomised controlled trials and their characteristics are shown in the Table 1, Table S1 and S2. In total, the trials included in the review reported ten different relevant symptoms. For each symptom, data could be expressed according to the following three outcomes: number of events, severity and time, as shown in Table S3.

**Table 1.**
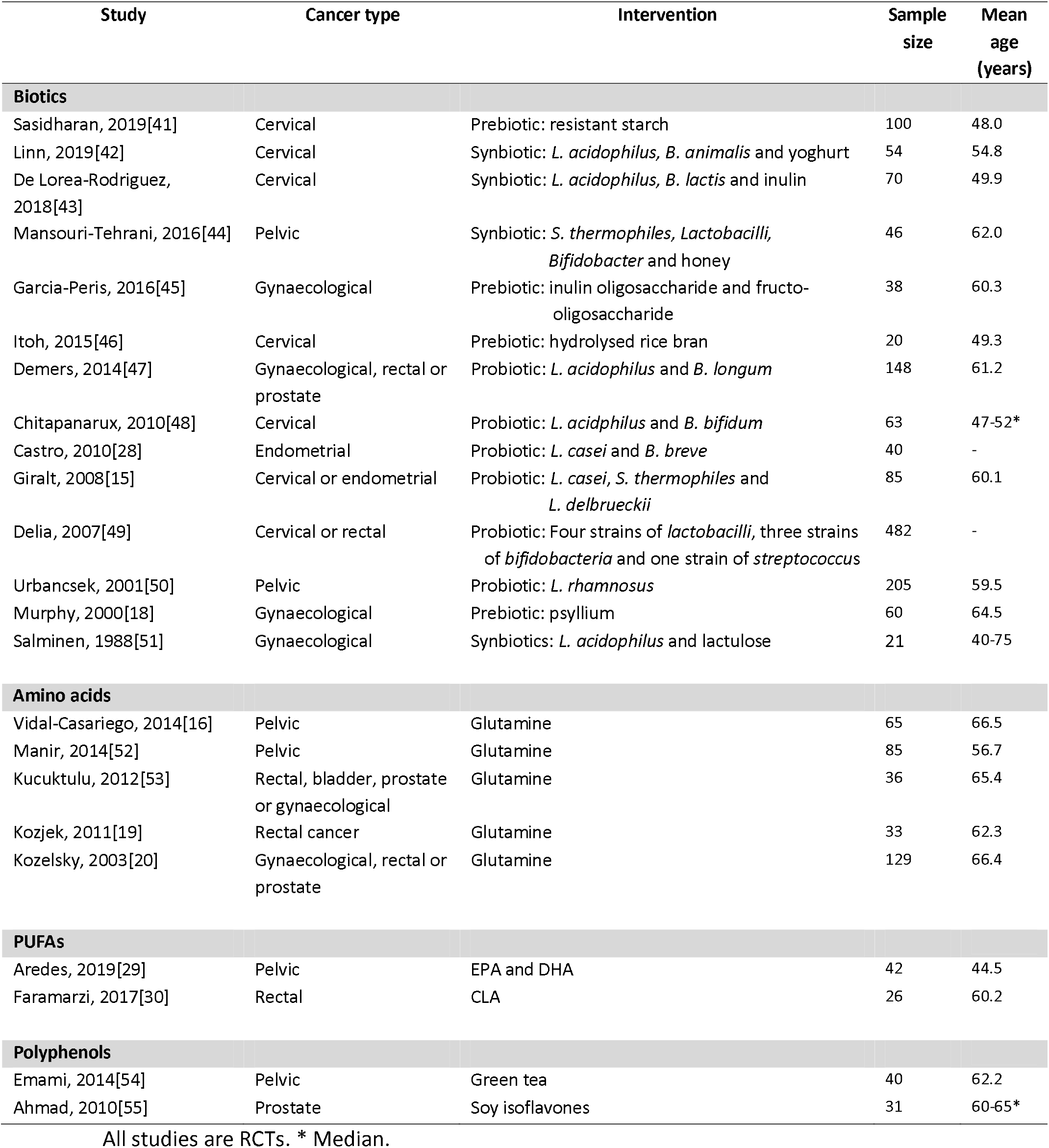
Characteristics of included studies.

### Efficacy of dietary supplements in preventing diarrhoea

The meta-analysis comprising 1,625 patients showed that dietary supplements reduced the risk of diarrhoea (Figure 3). The pooled risk ratio (RR) was 0.79 (95% CI: 0.66 to 0.94; P=0.007) and there was significant heterogeneity amongst the studies (I^2^=73%; P<0.001). The funnel plot for this meta-analysis (Figure S2) was largely symmetrical, implying only mild publication bias in the references included. There was no evidence that heterogeneity was due to mean age or sex of participants or sample size of the studies. Meta-analyses were then carried out for biotic, amino acid, poly-unsaturated fatty acid and polyphenol interventions.

**Figure 3.**
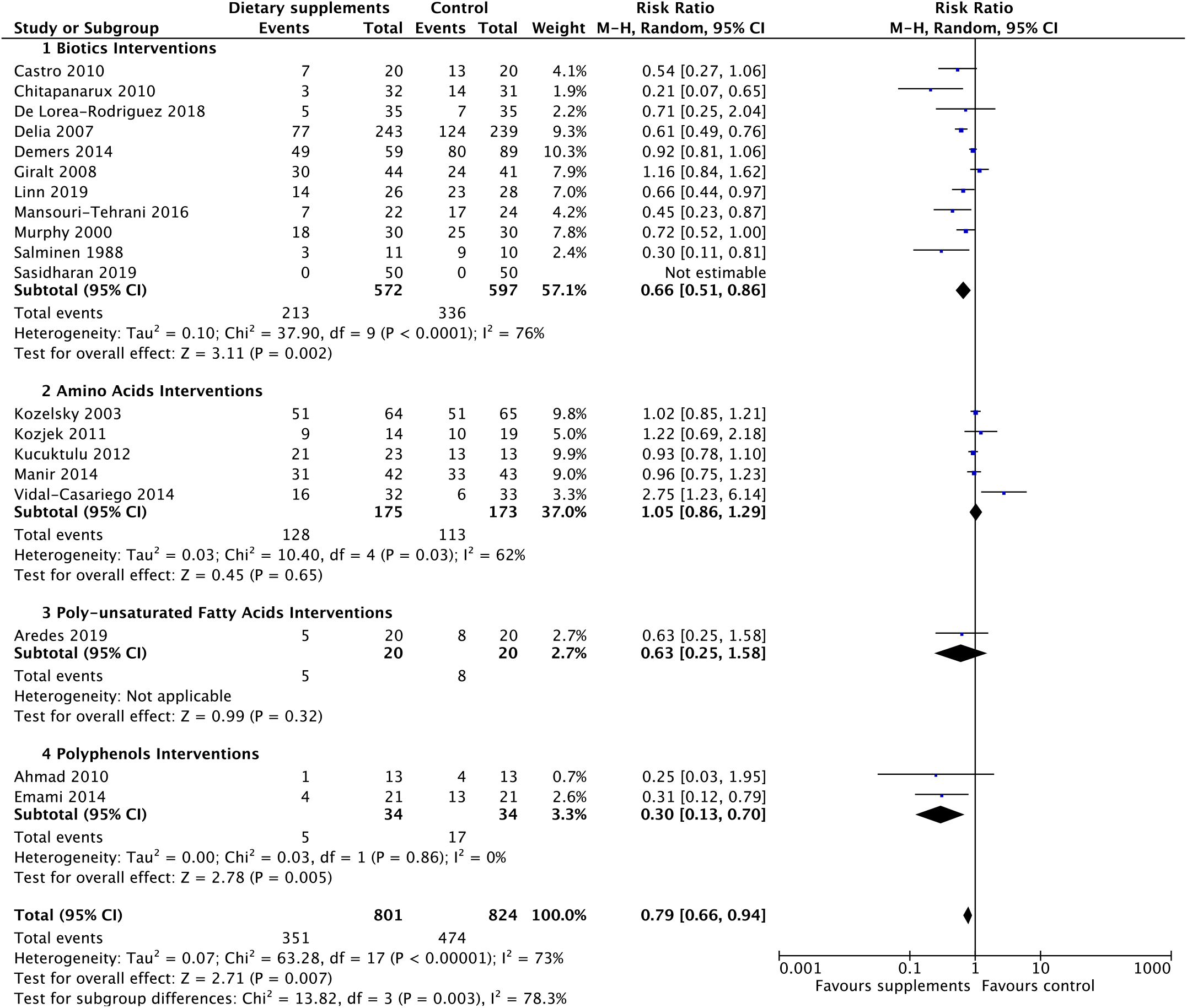
Forest plot of effects of biotic, amino acid, PUFA and polyphenol supplements on incidence of diarrhoea.

#### Efficacy of biotics in preventing diarrhoea

Biotic interventions significantly reduced the risk of diarrhoea with a RR of 0.66 (95% CI: 0.51 to 0.86; P=0.002) (Figure 3). All studies, except Giralt et al[15], had a RR of less than 1, suggesting the protective role of biotics against diarrhoea. The heterogeneity, I^2^, among these studies was 76% (P<0.001), so further analysis of the subclasses of probiotics and synbiotics was performed (Figure S3). The risk ratio for synbiotics (RR=0.45; 95% CI: 0.28 to 0.73) was lower than for probiotics (RR=0.71; 95% CI: 0.52 to 0.99). There was little heterogeneity among studies on synbiotics (I^2^=0%; P=0.50). Subgroup analysis was conducted by brachytherapy and chemotherapy (Figure S4 and S5). The results showed that patients not receiving brachytherapy (RR=0.63; 95% CI: 0.54 to 0.73) or not receiving chemotherapy (RR=0.62; 95% CI: 0.52 to 0.74) in particular benefited from probiotics and synbiotics.

#### Efficacy of amino acids in preventing diarrhoea

Amino acid interventions were not associated with risk of diarrhoea with a RR of 1.05 (95% CI=0.86 to 1.29; P=0.65, Figure 3). Five studies which included 348 patients were used to compare the incidence of diarrhoea between intervention and control groups. We found that four studies had consistent results of RR which were close to 1, but only Vidal-Cassariego et al reported a high RR of 2.75. There was high heterogeneity among studies (I^2^=62%, P=0.03)[16].

#### Efficacy of polyphenol in preventing diarrhoea

Two studies compared polyphenols and placebo among 64 patients (Figure 3). Both showed that the intervention was associated with lower incidence of diarrhoea. The overall RR was 0.30 (95% CI=0.13 to 0.70, P=0.005). There was no evidence of heterogeneity between these two studies (I^2^ = 0%, P=0.86).

### Efficacy of dietary supplements in preventing moderate to severe diarrhoea

Efficacy of dietary supplements was assessed against moderate to severe diarrhoea, with the incidence of moderate to severe diarrhoea defined as the incidence of grade 2 or higher diarrhoea, based on Common Terminology Criteria for Adverse Events (CTCAE) (older version: Common Toxicity Criteria)[17], except Murphy et al using the Murphy Diarrhoea Scale (MDS)[18] and Kozjek et al using their own criteria[19] (Figure 4). Dietary supplements were associated with a lower risk of moderate to severe diarrhoea (RR=0.68; 95% CI: 0.48 to 0.98; P=0.04). There was high heterogeneity among studies (I^2^=65%, P=0.009). The subgroup analysis suggested that the association was mainly driven by biotic interventions for which the RR was 0.49 (95% CI: 0.36 to 0.67; P<0.001), but not amino acids (RR=1.05; 95% CI: 0.82 to 1.34; P=0.70).

**Figure 4.**
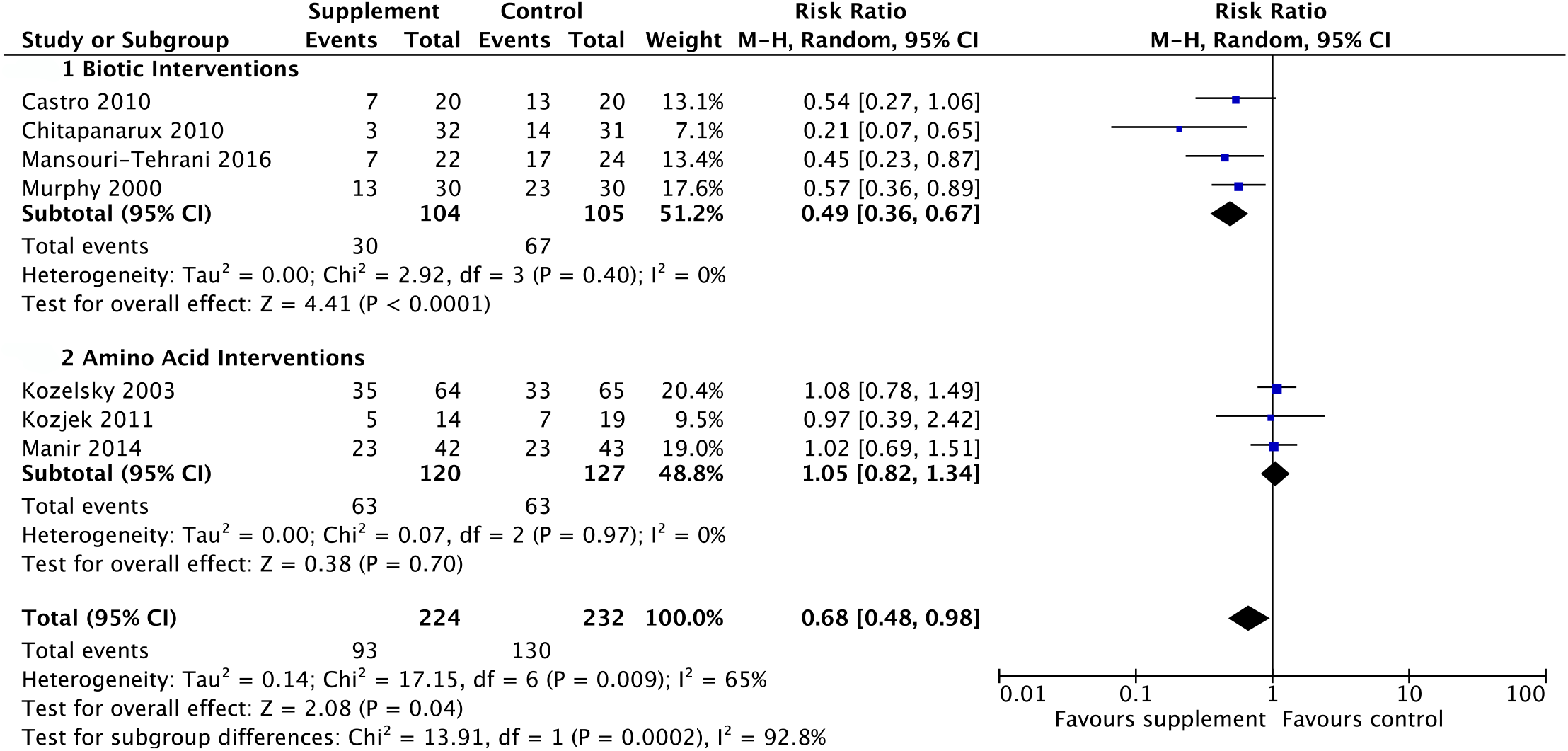
Forest plot of effect of dietary supplements on incidence of moderate to severe diarrhoea.

### Efficacy of dietary supplements in preventing the use of anti-diarrhoeal medication

Anti-diarrhoeal medication, such as loperamide, is often employed for patients who experience diarrhoea during or after radiotherapy. Therefore, we measured the effect of dietary supplements against the incidence of anti-diarrhoeal medication use (Figure 5), and found that dietary supplements were associated with lower risk of anti-diarrhoeal medication use in patients (RR=0.67; 95% CI: 0.47 to 0.96; P=0.03) and there was high heterogeneity among studies (I^2^=62%, P=0.01). For biotic interventions, the RR was 0.64 (95%CI: 0.44 to 0.92) and heterogeneity was intermediate (I^2^=45%; P=0.11).

**Figure 5.**
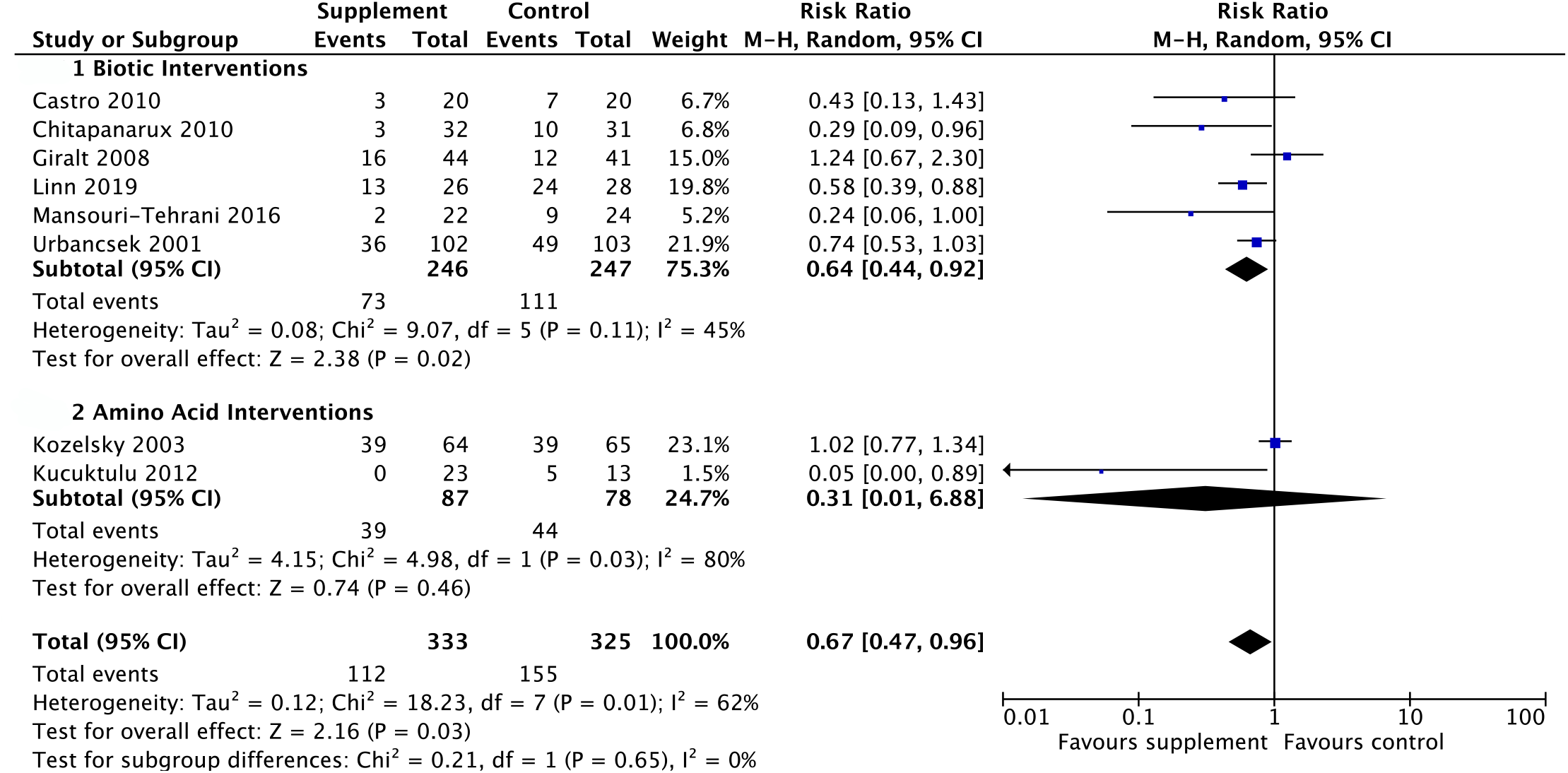
Forest plot of effect of dietary supplements on incidence of anti-diarrhoeal medication use.

### Effects of dietary supplements on nausea, vomiting, flatulence/bloating, bowel movement frequency, tenesmus and blood in bowel movement

Table S4 lists the results of meta-analysis of the other outcomes, including nausea, vomiting, flatulence/bloating and bowel movement frequency. Dietary supplements tended to decrease the risk of nausea (RR=0.74; 95% CI: 0.36 to 1.50; P=0.40) and the mean number of bowel movements per day (mean difference=-3.88; 95% CI: −10.29 to 2.52; P<0.001). The results also showed that the interventions had no effect on vomiting and flatulence/bloating with relative risks of 0.99 (95% CI: 0.79 to 1.25, P=0.95) and 1.12 (95% CI: 0.59 to 2.12; P=0.72) respectively. Only Kozelsky et al studied the outcomes of tenesmus and blood in bowel movements and found that glutamine had no effect on these symptoms[20].

## DISCUSSION

In this review, 23 randomised controlled trials met inclusion criteria for quantitative analysis. Risk of bias assessment was conducted for each of these studies. Meta-analyses were carried out for seven of these outcomes. These showed that dietary supplements are effective in reducing the risk of diarrhoea, experiencing moderate to severe diarrhoea and anti-diarrhoeal medication use. Subgroup analysis showed that biotic supplements and polyphenols were effective in reducing the risk of these outcomes, but amino acids were ineffective. Among the subclasses of biotic interventions, both probiotic and synbiotic supplements were shown to be effective in reducing the risk of diarrhoea, particularly among patients not receiving brachytherapy or chemotherapy (Figure S4A and S5A). Taken together, these results indicate that biotic supplements can reduce the risk of acute diarrhoea and the severity of diarrhoea in patients undergoing pelvic radiotherapy.

Our study is the only reported meta-analysis that investigates the efficacy of four different classes of dietary supplements on acute symptoms of gastrointestinal toxicity. Most previous meta-analyses investigated only a single class of dietary supplement. This review includes more studies (23) and patients (1,919) than other published reviews, increasing confidence in the quantitative analysis. Several meta-analyses have been conducted for probiotic and synbiotic supplements over the last decade, but the category of prebiotics and subgroups of patients receiving brachytherapy or chemotherapy have not yet been analysed. Therefore, the inclusion of prebiotics in this review is another key distinguishing feature.

The previous meta-analyses investigating the effects of biotic supplements on acute symptoms of gastrointestinal toxicity are listed in Table S5. Overall, the trials included in each meta-analysis are largely identical. Where different selections were made, this was mainly due to variations in search strategy and the statistical methods used. A Cochrane systematic review has also investigated the efficacy of interventions, including radiotherapy techniques and pharmacological and non-pharmacological interventions, on acute and late adverse gastrointestinal effects of pelvic radiotherapy for primary pelvic cancers[4]. Compared to their study of non-pharmacological interventions, including dietary interventions, probiotics, glutamine, counselling, and protein supplements, our focused approach showed that the beneficial effects mainly came from the probiotics and synbiotics. Also, the focus of the Cochrane review was on prevention, rather than treatment, of acute symptoms of gastrointestinal toxicity, an important difference to our review. Additionally, their search was only updated to November 2017; our search to June 2020 included three more recent studies of biotic supplements (224 patients) and one study focusing on PUFA supplements (40 patients). Currently, there are no published meta-analyses that investigate the effect of PUFA or polyphenol supplements on acute symptoms of gastrointestinal toxicity, and two included studies of polyphenols suggested that they are beneficial in preventing diarrhoea. One analysis investigating amino acid supplements (glutamine) was identified[21] but rejected because of different endpoints used. The authors included 13 RCTs investigating acute symptoms of radiation enteritis, with the following outcomes, abdominal cramping, blood in bowel movement and tenesmus. The researchers found that amino acid supplements were not effective in improving symptoms. This finding is consistent with the result of this review that amino acid supplements are ineffective in improving acute symptoms of gastrointestinal toxicity.

A strength of our meta-analysis is that the data relate to patients undergoing pelvic radiotherapy. They are not derived from cell lines or animal model systems. A further strength is that the trials are based on direct measurements of the symptoms of gastrointestinal toxicity, rather than less direct measures of effect, such as immunological, biochemical or histological markers. Therefore, the findings of this review can be directly applied to clinical practice. However, patient-related factors of low numbers (<40) of participants in 26% of included studies represent a weakness in the statistical treatment of data sets, resulting in wider confidence intervals. Most data were derived from single-centre trials, thus limiting the ability to recruit larger numbers of patients.

Furthermore, the baseline heterogeneity of the patient cohorts is high, as it is not possible to subdivide patients into meaningfully representative sub-groups in terms of sex, lifestyle-related factors, co-morbidities and type of cancer. To minimise the confounding effects of heterogeneity, we performed meta-regressions of age and radiation dose on the incidence of diarrhoea (Figure S1) and subgroup analyses by patients receiving brachytherapy (Figure S4) or chemotherapy (Figure S5). We did this because studies showed that the risk factors for radiation enteritis include older age[5], the dose of radiation used[22], combining internal and external RT[23] and the concomitant use of chemotherapy[24]. Figure S1 show that the effects of interventions on incidence of diarrhoea did not vary by mean age (A) and RT dose (D). It is also noted that the RT doses and techniques of most of the studies were homogenous. Seventeen out of 23 studies used an RT dose of approximately 50 Gy, the exceptions being Delia et al[25], Murphy et al[26], Ahmad et al[27] which used differing higher doses, and 3 studies that did not specify the dosage, including Castro et al[28], Aredes et al[29] and Faramarzi et al[30]. Almost all studies used conventional external beam radiotherapy, except Ahmad et al[27] using 3D conformal radiation and IMRT (Table S1). To measure the incidence of diarrhoea, 13 studies used the scale of Common Toxicity Criteria (CTC) or Common Terminology Criteria for Adverse Events (CTCAE). The other studies used either Bristol stool form scale (BSFS), WHO toxicity grading, Radiation Therapy Oncology Group (RTOG) toxicity scale, Murphy Diarrhea Scale (MDS), adapted NCI questionnaire, European Organization for Research and Treatment of Cancer Quality of Life Questionnaire version.3.0 (EORTC QLQ-C30) or non-specified Quality of life (QOL) questionnaire. These have similar definitions of diarrhoea, which enabled us to combine these trials (Table S2). The heterogeneity may reduce the evidence certainty of this study, so future methodologically well-designed, large-scale trials are needed to strengthen the evidence of the benefits of dietary supplements.

The underlying protective effects of dietary supplements against GI toxicities may be mediated as shown in Figure S6. A direct effect on the intestinal immune environment following intake of specific dietary agents may lead to anti-inflammatory changes that alleviate gastrointestinal toxicity. There may also be an indirect effect, whereby the above immunomodulatory actions are developed in response to changes in the gut microbiota and their metabolites, particularly SCFAs.

In terms of probiotics, these can act in three beneficial ways, including down-regulation of epithelial apoptosis and intestinal inflammatory activity of the digestive tract[31], production of the SCFA butyrate[32] and provision of resistance against infection of the damaged mucosa by pathogens[33]. Having both properties of prebiotics and probiotics, synbiotic may confer a superior effect[34].

This review aimed to investigate the effect of dietary supplements on acute and late symptoms, but there were no studies reporting on late side effects among included studies. Chronic symptoms of gastrointestinal toxicity typically emerge a few months to years following irradiation and occur in most of the intestinal compartments[35]. The prevalence of chronic symptoms is reported to be between 5 and 15%[36] and can result in potentially fatal complications such as fistulation, sepsis and perforation of the colon. Due to improvements in pelvic cancer treatment, the number of cancer survivors is increasing. Consequently, more patients are potentially at risk of chronic symptoms of gastrointestinal toxicity. However, improvements in pelvic cancer treatment, such as the use of image guided radiotherapy, have in turn resulted in a decrease in chronic radiation effects[37]. Evidence from clinical studies suggests that acute and chronic effects are linked, with the risk of developing late effects greater in patients that have developed acute effects (consequential late effects)[38, 39]. Consistent with this finding, a systematic review that investigated the relationship between acute and late gastrointestinal toxicity after RT for prostate cancer concluded that acute GI toxicity may be predictive of high risk of developing late effects[40].

## CONCLUSION

Findings from our systematic review and meta-analysis suggest that biotic supplements, specifically probiotics and synbiotics, are effective in reducing the risk and severity of acute symptoms of gastrointestinal toxicity caused by pelvic radiotherapy, particularly among patients not receiving brachytherapy or chemotherapy. The data also broadly confirm that they are safe to use.

## Data Availability

N/A

## Acknowledgements

N/A

