## Supplementary data for "Interventions with dietary supplements, including pre-, pro- and synbiotics, to reduce acute and late gastrointestinal side effects in patients undergoing pelvic radiotherapy: A systematic review and meta-analysis"

Benjamin Bartsch^1#^, BSc, Chee Kin Then^1#^, MD, MRes, Elinor Harriss^2^, MA, MSc, MSc, Christiana Kartsonaki^3,4^, MSc, DPhil, Anne E. Kiltie^1^*, MA, DM, DSc, MRCP, FRCR

^1^Oxford Institute for Radiation Oncology, Department of Oncology, University of Oxford, Oxford, UK

^2^Bodleian Health Care Libraries, University of Oxford, Oxford, UK

^3^Clinical Trial Service Unit & Epidemiological Studies Unit (CTSU), Nuffield Department of Population Health, University of Oxford, Oxford, UK

^4^Medical Research Council Population Health Research Unit (MRC PHRU) at the University of Oxford, Nuffield Department of Population Health, University of Oxford, Oxford, UK

#These authors contributed equally to this work.

***Corresponding author name & email address**

Professor Anne E Kiltie. Oxford Institute for Radiation Oncology, Department of Oncology, University of Oxford, Old Road Campus Research Building, Off Roosevelt Drive, Oxford OX3 7DQ; Phone: +44 1865 617352;

**Author responsible for statistical analysis name & email address**

Dr. Christiana Kartsonaki.

**Supplementary data**

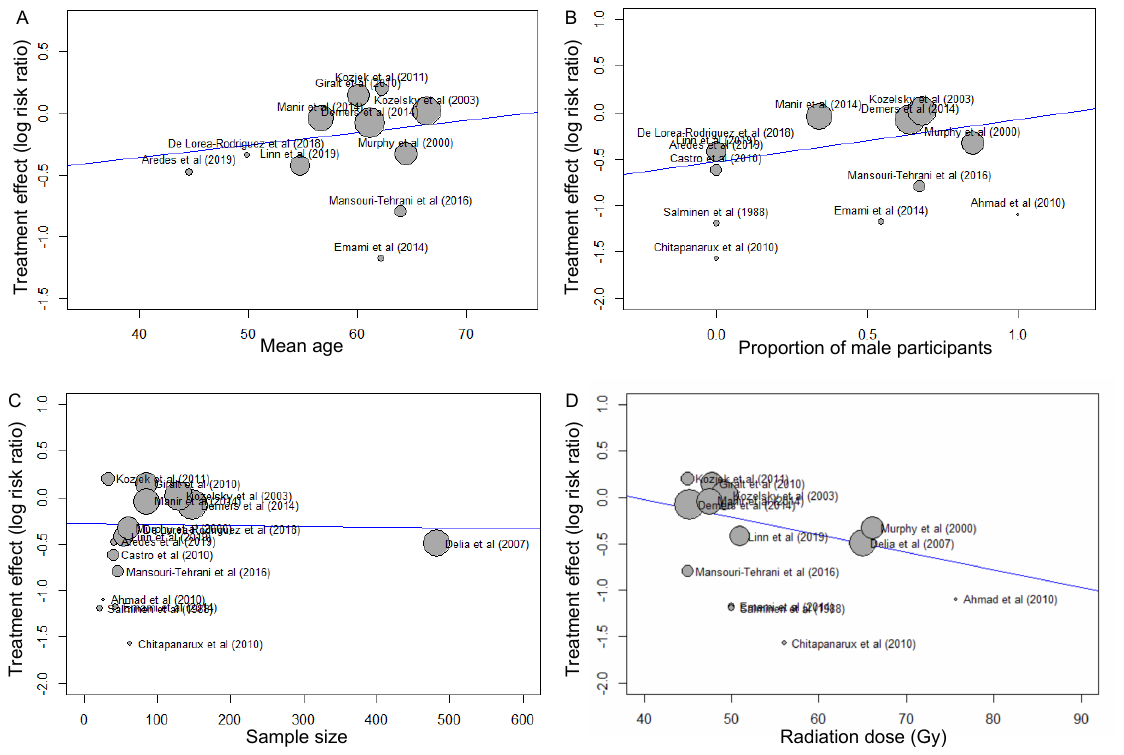

**Figure S1** Meta-regression by mean age, proportion of male participants, sample size and RT dose to the effects of interventions

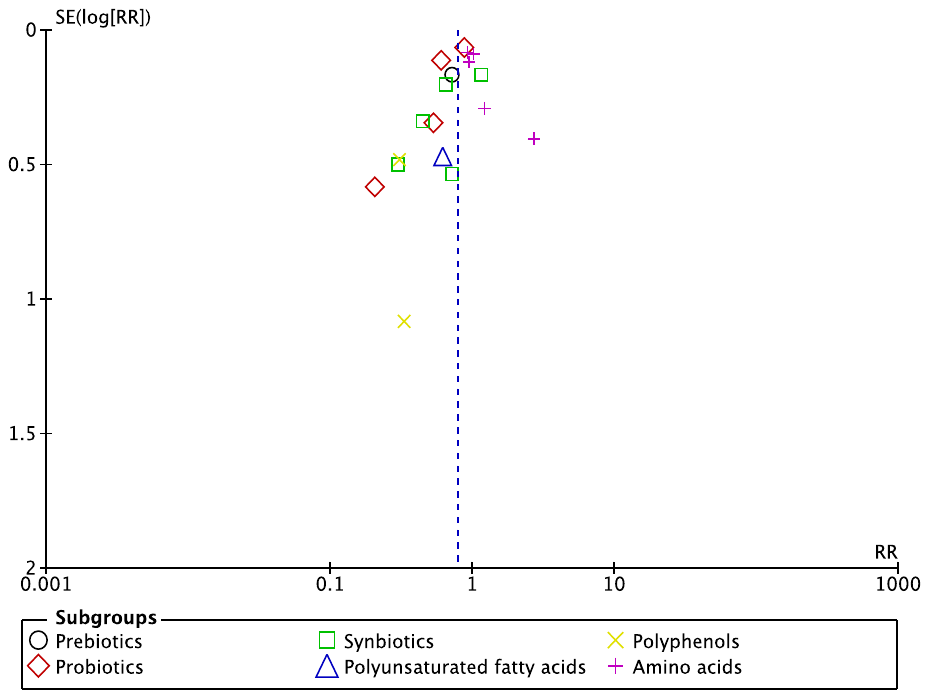

**Figure S2** Funnel plot for the meta-analysis that investigated the effect of dietary supplements on incidence of diarrhoea.

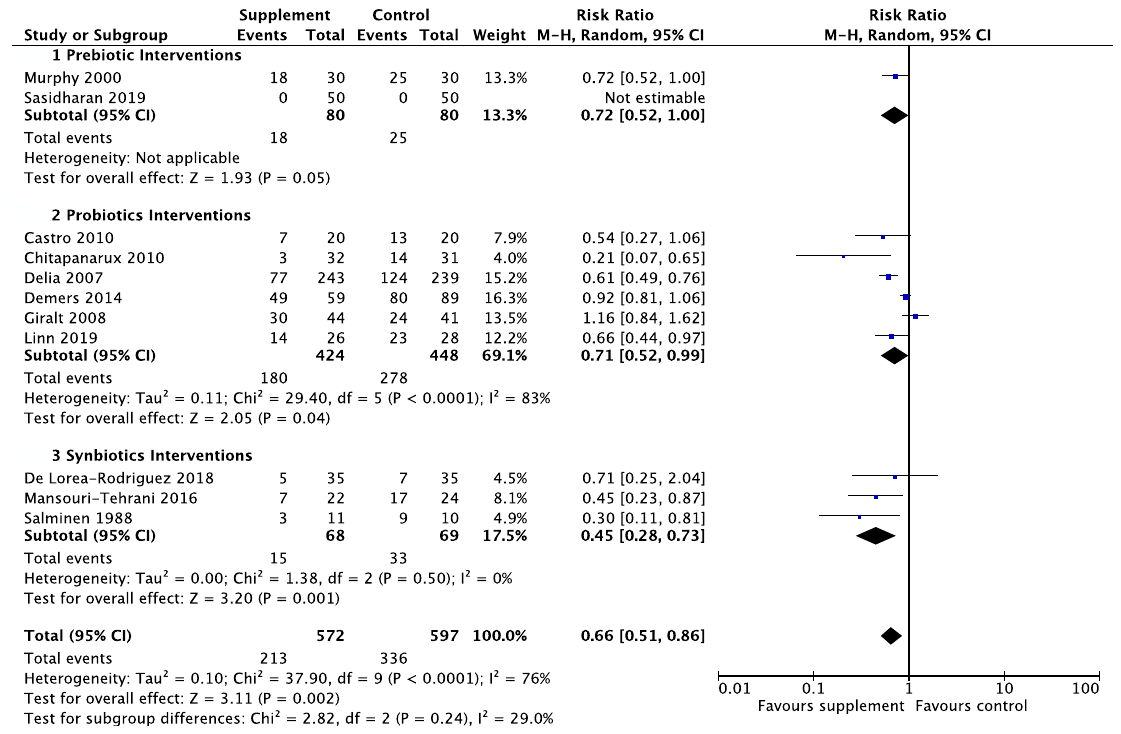

**Figure S3** Forest plot of effect of prebiotic, probiotic and synbiotic supplements on incidence of diarrhoea.

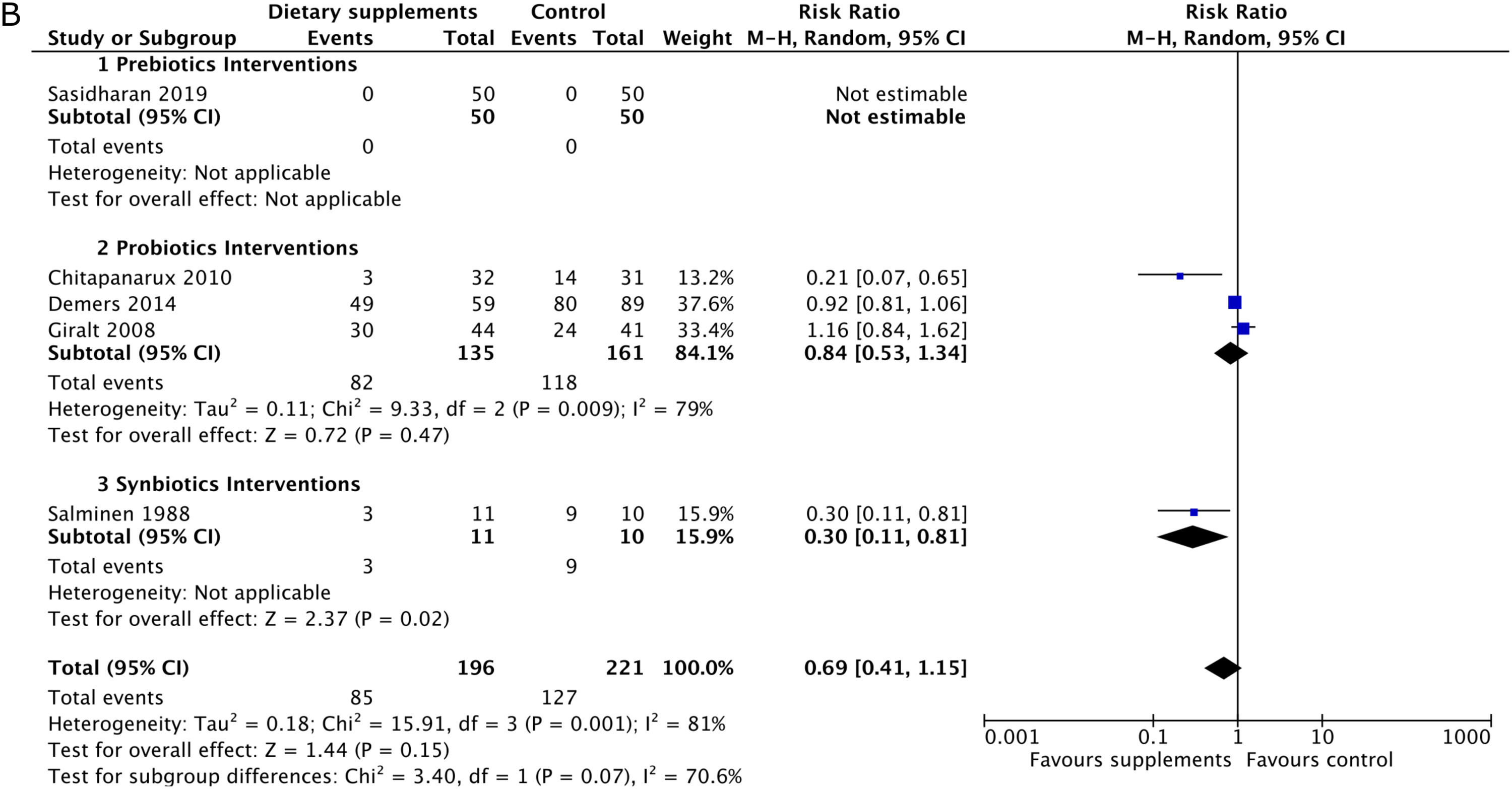

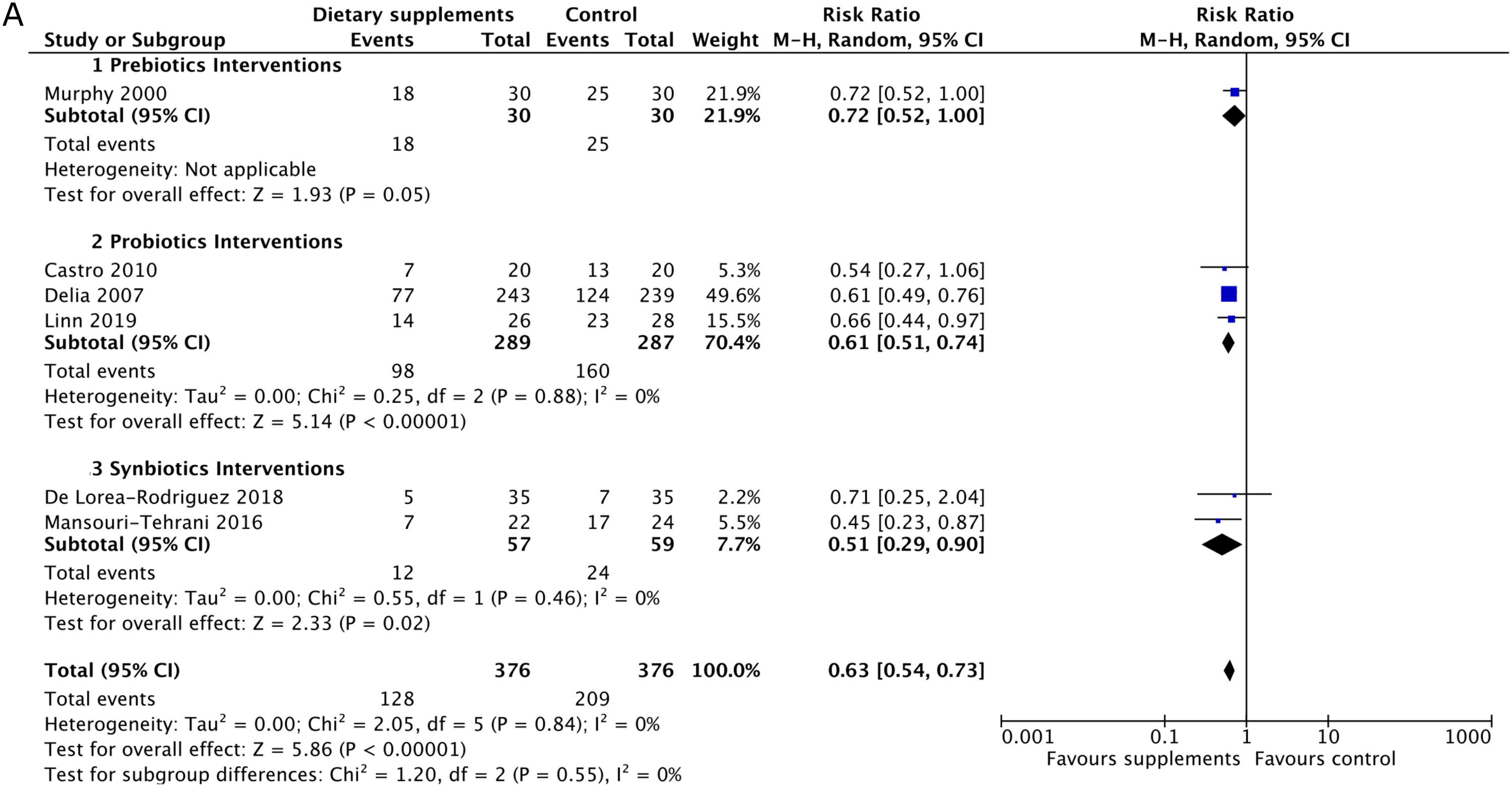

**Figure S4** Subgroup analysis of effect of prebiotic, probiotic and synbiotic supplements on incidence of diarrhoea by brachytherapy.

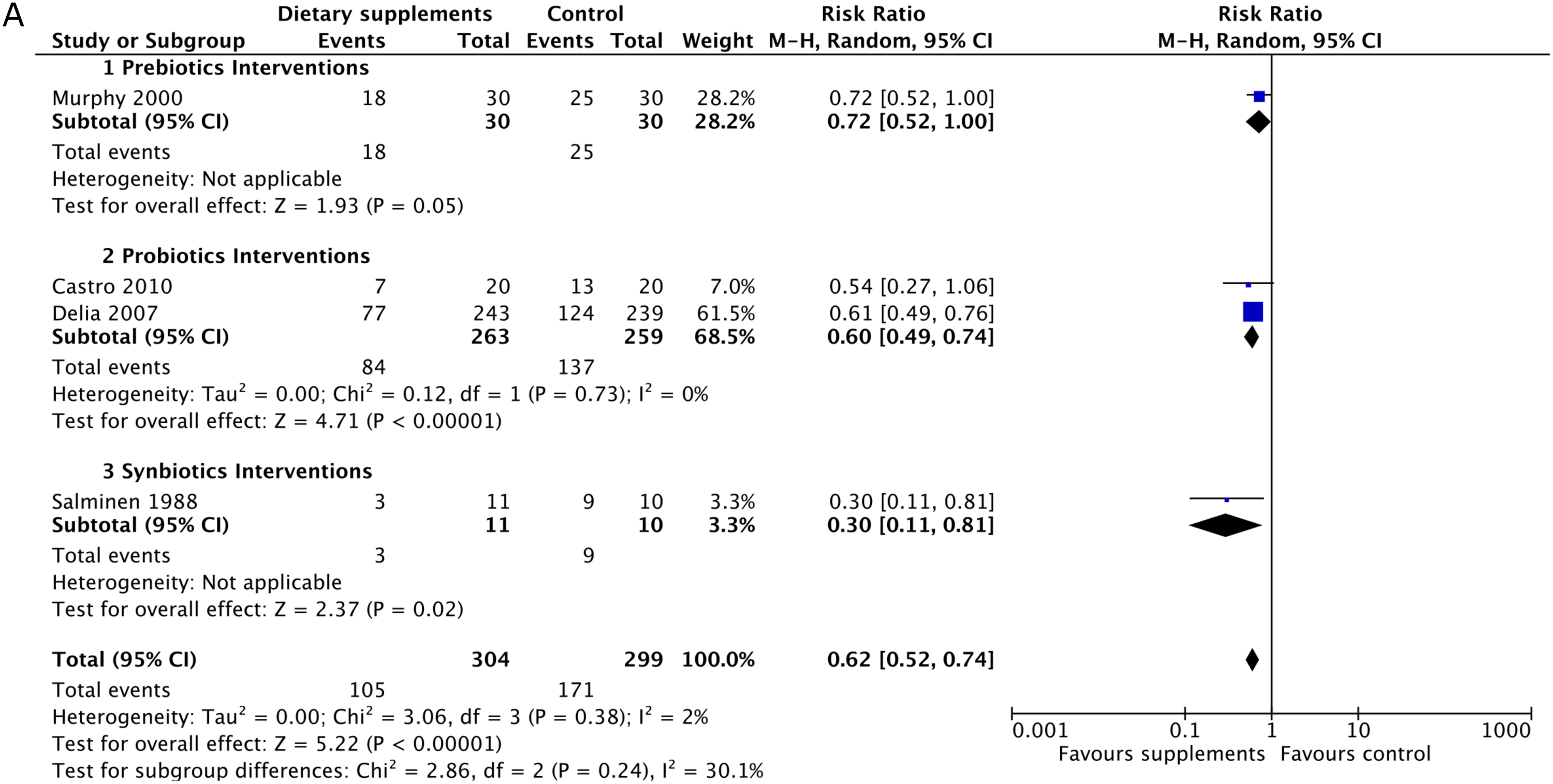

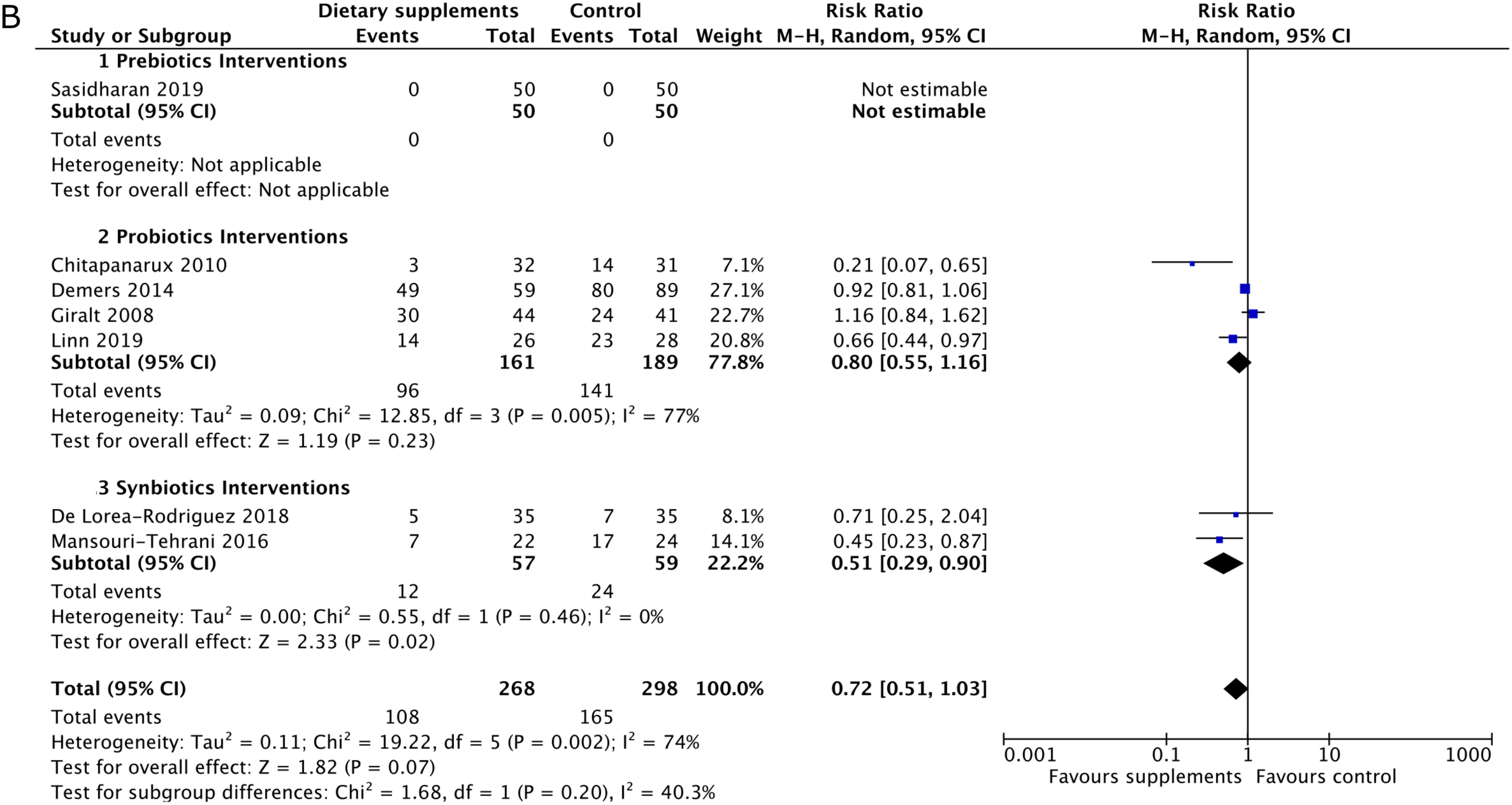

**Figure S5** Subgroup analysis of effect of prebiotic, probiotic and synbiotic supplements on incidence of diarrhoea by chemotherapy.

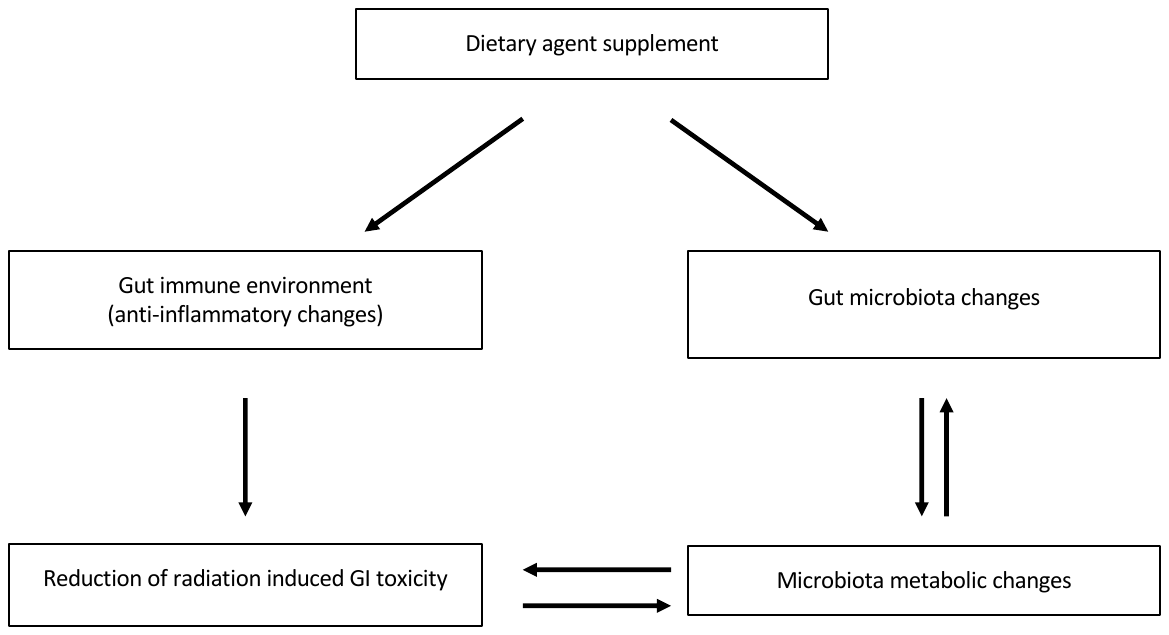

**Figure S6** Schematic overview of potential benefits of dietary supplement intervention to alleviate symptoms of gastrointestinal toxicity caused by pelvic radiotherapy.

**Table S1 Details of radiotherapy, brachytherapy and chemotherapy**

| **Study** | **Type of treatment** | **Mean of radiotherapy dose (technique)** | **Brachytherapy** | **Chemotherapy**  **(% of patients or type)** |
| --- | --- | --- | --- | --- |
| **Biotics** |  |  |  |  |
| Sasidharan | Radical chemoradiotherapy | 50 Gy in 25 fractions  (cobalt 60 gamma rays or 6 MV/15 MV beams from a linear accelerator) | Yes | Cisplatin |
| Linn | RT with/without chemotherapy | 51 Gy in 25 fractions  (2D RT with linear accelerator) | - | 75.9% |
| De Lorea-Rodriguez | Chemoradiotherapy | N/S | - | N/S |
| Mansouri-Tehrani | RT or chemoradiotherapy | 40 - 50 Gy and 1.8 Gy/fraction  (conventional RT with 18 MV) | - | 38.8% |
| Garcia-Peris | Post-operative RT | 52.2 Gy and 1.8 Gy/fraction  (linear accelerator with 15 MV) | Yes | Excluded |
| Itoh | Chemoradiotherapy | 50.4 Gy in 28 fractions  (external beam RT) | Yes | Cisplatin and 5-FU |
| Demers | Post-operative RT or chemoradiotherapy | 40-50.4 Gy and 1.8-2.0 Gy/fraction (external RT) | Some | 52.4%; cisplatin (cervical cancers); 5-FU or capecitabine (rectal cancers) |
| Chitapanarux | Chemoradiotherapy | 56 Gy and 2 Gy/fraction  (external beam RT) | Iridium-192 | Cisplatin |
| Castro | Pelvic RT | N/S | - | - |
| Giralt | Postoperative pelvic RT or chemoradiotherapy | 45–50.4 Gy and 1.8–2 Gy/fraction  (linear accelerator with 15-18 MV) | Yes | Cisplatin (cervical carcinoma) |
| Delia | Postoperative pelvic RT | 60-70 Gy (X-ray) | - | - |
| Urbancsek | Lower abdomen RT | 50 Gy* and 2 Gy/fraction | - | - |
| Murphy | Pelvic RT | 66.1 Gy in 20 fractions | - | - |
| Salminen | Post-operative internal and external pelvic RT | 50 Gy for the pelvic area,  80 Gy for the tumour in 22 fractions | Intracavitary caesium | - |
| **Amino acids** |  |  |  |  |
| Vidal-Casariego | Abdominal and pelvic RT | 50.7 Gy and 1.8 - 1.9 Gy/session | 24.6% | 40.5% |
| Manir | Pelvic RT with/without chemotherapy | 45 - 50 Gy and 2 Gy/fraction  (Cobalt 60 machine) | - | 84.7% |
| Kucuktulu | Pelvic RT | 45–50.4 Gy in 1.8–2.0 Gy fractions for whole pelvis  45–70 Gy for planned RT | - | 5-FU (rectum carcinoma), cisplatin (bladder and gynecologic cancers) |
| Kozjek | Pre-operative chemoradiotherapy | 45 Gy to the pelvis 1.8 Gy/fractions and 5.4 Gy as a boost to the primary tumour | - | Capecitabine |
| Kozelsky | Pelvic RT | 45 to 53.5 Gy and 1.7-2.1/fraction for pelvis | - | 6.9%; 5-FU |
| **PUFAs** |  |  |  |  |
| Aredes | Chemoradiotherapy | N/S | - | Cisplatin-based chemotherapy |
| Faramarzi | Preoperative chemoradiotherapy | N/S | - | N/S |
| **Polyphenols** |  |  |  |  |
| Emami | Abdominal and pelvic RT | 50 Gy and 10 Gy weekly | - | 35.7% |
| Ahmad | Curative RT | 73.8 to 77.5 Gy and 1.8-2.5 Gy/fraction  (conformal and/or IMRT) | - | Excluded |

* Median. Abbreviations: 5-fluorouracil (5-FU), radiotherapy (RT), not specified (N/S)

**Table S2 Details of the dietary supplements used and outcome severity of diarrhoea**

|  | **Intervention** | | | **Outcome of diarrhoea** | |
| --- | --- | --- | --- | --- | --- |
| **Study** | **Dose** | **Usage** | **Timing** | **Scale** | **Grade; timepoint** |
| **Biotics** |  |  |  |  |  |
| Sasidharan | 30 g high amylose corn starch containing 72% amylose | b.i.d | During 6-week RT period only | CTC | ≥grade 2;  at 6^th^ week of RT |
| Linn | 300mg functional yogurt containing  1.75 x 10^9^ CFU *L. acidophilus* LA-5 plus *B. animalis* subsp. *Lactis* BB-12 | t.i.d | During RT period only | CTCAE | ≥grade 1;  weekly for 3 weeks after RT completion |
| De Lorea-Rodriguez | Blue agave inulin,  2 x 10^8^ CFU *L. acidophilus* NCFM,  2 x 10^7^ CFU *B. lactis* Bi-07 | t.i.d | 7 weeks | BSFS | Type 6,7;  at 7^th^ week |
| Mansouri-Tehrani | 15 mg honey,  1.5 x 10^9^ CFU *L. casei*, 1.5 x 10^10^ CFU *L. acidophilus*,  3.5 x 10^9^ CFU *L. rhamnosus*, 2.5 x 10^8^ CFU *L. bulgaricus*,  1 x 10^10^ CFU *B. breve*, 5 x 10^8^ CFU *B. longum*,  1.5 x 10^8^ CFU *Streptococcus thermophilus* per 500 mg | b.i.d | 1 week before RT and continued for 4-week RT period | CTC | Grade 2&3;  during RT period |
| Garcia-Peris | 6 g fibre mixture containing 50% inulin and 50% FOS | b.i.d | 1 week before until 3 weeks after RT | CTC | - |
| Itoh | 1 g hydrolysed rice bran | t.i.d | 1 week before until end of RT | CTCAE | - |
| Demers | 1.3 x 10^9^ (standard) / 1 x 10^10^ (high) CFU *L. acidophilus* LAC-361 plus *B. longum* BB-536 | b.i.d/  t.i.d | During RT period only | WHO toxicity criteria | ≥grade 2;  at 60^th^ day |
| Chitapanarux | 2 x 10^9^ units of a *L. acidophilus* plus *B. bifidum* | b.i.d | 1 week before until end of RT | CTC | ≥grade 2;  at RT completion |
| Castro | *L. casei* and *B. breve* | N/S | N/S | CTC | ≥grade 2; N/S |
| Giralt | 96 mL of a fermented liquid yogurt containing around  10^8^ CFU/g of *L. casei* DN-114 001, *Streptococcus thermophilus* and *L. delbrueckii,* subsp. *Bulgaricus* | t.i.d | 1 week before until end of RT | CTC | ≥grade 2;  at RT completion |
| Delia | 450 x 10^9^ CFU viable lyophilized bacteria /g including  *L. casei, L. plantarum, L. acidophilus, L. delbruekii* subsp. *Bulgaricus,* *B. longum, B. breve, B. infantis*, *Streptococcus salivariu*s subsp. *thermophilus* | t.i.d | During RT period only | WHO toxicity grading | N/S;  weekly during RT until 1 month after RT |
| Urbancsek | 1.5 x 10^9^ CFU *L. rhamnosus* | t.i.d | - | - | - |
| Murphy | Psyllium | - | - | MDS | N/S; weekly during RT until 1 month after RT |
| Salminen | 150 ml of a fermented milk test product with at least  2 × 10^9^ CFU *L. acidophilus* bacteria and 6.5% lactulose | q.d | 5 days before until 10 days after RT | - | N/S;  6 weeks after treatment |
| **Amino acids** |  |  |  |  |  |
| Vidal-Casariego | 30 g glutamine | q.d | 3 days before until end of RT | RTOG toxicity scale | Soft or liquid stools;  during 2^nd^ visit |
| Manir | 10 g glutamine | q.d | During 5-week RT period only | CTCAE | ≥grade 1;  weekly during RT until 1 week after RT |
| Kucuktulu | 15 g glutamine | t.i.d | 1 week before until 1 week after RT | CTC | ≥grade 1; N/S |
| Kozjek | 10 g glutamine | t.i.d | During 5-week RT period only | Adapted NCI questionnaire | ≥grade 2;  at RT completion |
| Kozelsky | 4 g glutamine | b.i.d | Start of RT until 2 weeks after RT | CTC | ≥grade 1; N/S |
| **PUFAs** |  |  |  |  |  |
| Aredes | 2 g eicosapentaenoic acid and  450 mg docosahexaenoic acid | q.d | For 45 days, corresponding to duration of chemoradiotherapy | CTCAE | ≥grade 2; at end of chemoradiotherapy |
| Faramarzi | 3 g conjugated linoleic acid / capsule  (1 capsule at breakfast and dinner, 2 capsules at lunch) | t.i.d | 1 week until end of chemoradiotherapy | EORTC QLQ-C30 | - |
| **Polyphenols** |  |  |  |  |  |
| Emami | 450 g green tea | q.d | During 5-week RT period only | CTC | Grade 1;  at 5^th^ week |
| Ahmad | 100 mg soy isoflavones | b.i.d | For 6 months beginning with the first day of RT | QOL questionnaire | N/S; 6 months from the start of RT |

The route for all studies was oral. Abbreviations: colony-forming unit (CFU), Not specified (N/S), everyday (q.d), two time a day (b.i.d), three times a day (t.i.d), Common Toxicity Criteria (CTC), Common Terminology Criteria for Adverse Events (CTCAE), Bristol stool form scale (BSFS), Murphy Diarrhea Scale (MDS), Radiation Therapy Oncology Group (RTOG), European Organization for Research and Treatment of Cancer Quality of Life Questionnaire version.3.0 (EORTC QLQ-C30), Quality of life (QOL).

**Table S3 Summary of key symptoms and outcomes**

| **Symptom** | **Outcomes** | |
| --- | --- | --- |
| - Diarrhoea - Stool consistency - Constipation - Need for anti-diarrhoeal medication - Bowel movements - Nausea - Vomiting - Abdominal pain - Appetite loss - Flatulence /bloating - Tenesmus - Rectal bleeding | Number of events | Incidence of events |
|  |  | Mean/median number of events |
|  | Severity | Incidence of specific severity grade(s) |
|  |  | Mean/median grade |
|  | Time | Mean/median time of onset |
|  |  | Mean/median time of duration |

Table S4 Meta-analyses of other gastrointestinal outcomes

| **Outcome** | **Number of studies** | **Number of participants** | **Risk ratio**  **(95% CI)** | **Test for**  **overall effect, P** | **Heterogeneity**  **I^2^ (%; P value)** |
| --- | --- | --- | --- | --- | --- |
| Incidence of vomiting | 4 | 237 | 0.99 (0.79, 1.25) | 0.95 | 59 (0.52) |
| Incidence of nausea | 3 | 192 | 0.74 (0.36, 1.50) | 0.40 | 0 (0.08) |
| Incidence of flatulence/bloating | 3 | 152 | 1.12 (0.59, 2.12) | 0.72 | 76 (0.01) |
| The mean number of bowel movements per day* | 3 | 1045 | -3.88 (-10.29, 2.52) | 0.23 | 99 (<0.01) |

*The outcome was mean difference.

Table S5 Comparison of this review with precedent meta-analyses that investigate the effect of biotic supplements on acute symptoms of gastrointestinal toxicity

| **Meta-analysis** | **Number of studies (patients no.)** | **Data overlap*** | **Differences in scope** | **Selected key findings** |
| --- | --- | --- | --- | --- |
| This review  (biotics only) | 14 (1,432) | - | Includes probiotics, prebiotics and synbiotics | Probiotics and synbiotics reduce the risk of radiation induced diarrhoea.  Synbiotics had greater efficacy.  Biotics as a whole reduce the risk of severity diarrhoea. |
| Qiu *et al*^1^ | 9 (1,508) | 8 out of 9 | Probiotics, focus on cervical cancer | Probiotics reduce the risk of radiation induced diarrhoea and the risk of severe diarrhoea. |
| Wei *et al*^2^ | 12 (1,554) | 8 out of 12 | Probiotics, also looks at chemotherapy | Researchers could neither demonstrate nor refute a beneficial effect of probiotics on symptoms of GI toxicity. |
| Liu *et al*^3^ | 6 (917) | 6 out of 6 | Probiotics | Probiotics reduce the risk of radiation induced diarrhoea. |
| Hamad *et al*^4^ | 6 (904) | 6 out of 6 | Probiotics | Probiotics reduce the odds of developing radiation induced diarrhoea. |
| Fuccio *et al*^5^ | 4 (632) | 4 out of 4 | Probiotics | Probiotics reduce the odds of developing radiation induced diarrhoea. |

*Proportion of studies overlapping with trials used in our meta-analysis. For example, among nine trials used in the study of Qiu *et al*, eight of them had been included in our meta-analysis.

**Appendices**

**Appendix 1. MEDLINE search strategy**

1	Pelvic Neoplasms/

2	exp Uterine Neoplasms/

3	Urinary Bladder Neoplasms/

4	Prostatic Neoplasms/

5	exp Rectal Neoplasms/

6	((pelvi* or cervi* or uter* or endometr* or bladder* or prostat* or rect* or anal or anus or genitourin* or genito-urin* or colorect* or gynaeco* or gyneco*) adj5 (cancer* or tumor* or tumour* or neoplas* or malignan* or carcinoma* or adenocarcinoma*)).mp.

7	1 or 2 or 3 or 4 or 5 or 6

8	exp radiotherapy/

9	radiotherapy.fs.

10	(radiotherap* or brachytherap* or radiat* or irradiat* or chemoradiotherap* or chemoradiat*).mp.

11	8 or 9 or 10

12	exp Probiotic/

13	exp Synbiotic/

14	exp Bacteria/

15	exp Prebiotics/

16	exp Diet/

17	Food Habits/

18	exp Food/

19	exp Nutrition Therapy/

20	exp Nutrition Disorders/

21	exp Feeding Methods/

22	exp Polysaccharides/ or exp Starch/

23	exp Amino Acids/

24	exp Peptides/

25	exp Antioxidants/

26	exp Fish Oils/

27	(nutrition* or nutrient* or malnutrition or diet* or food* or feed* or drink* or supplement* or formula* or vitamin* or mineral* or fat* or carbohydrate* or protein* or probiotic* or synbiotic* or bacteria* or prebiotic* or "amino acid*" or peptide*).mp.

28	(lactose or fibre or fiber or bran or psyllium or plantago ovata or ispaghula or polysaccharide* or polymeric* or peptide* or amino-acid* or glutamine* or fish oil* or arginine* or antioxidant* or anti-oxidant* or Metamucil or Inulin* or fructose-oligosaccharide* or fructo-oligosaccharide* or oligosaccharide* or starch or saccharide*).mp.

29	(enteral* or nasogastric* or gastrostomy of jejunostomy or oral* or tube* or sip*).mp.

30	12 or 13 or 14 or 14 or 15 or 16 or 17 or 18 or 19 or 20 or 21 or 22 or 23 or 24 or 25 or 26 or 27 or 28 or 29

31	randomized controlled trial.pt.

32	controlled clinical trial.pt.

33	randomized.ab.

34	placebo.ab.

35	clinical trials as topic.sh.

36	randomly.ab.

37	trial.ti.

38	exp Cohort Studies/

39	(cohort* or prospective* or retrospective*).mp.

40	Comparative Study/

41	(observational or comparative).mp.

42	(case* adj5 series).mp.

43	31 or 32 or 33 or 34 or 35 or 36 or 37 or 38 or 39 or 40 or 41 or 42

44	7 and 11 and 30 and 43

45	exp animals/ not humans.sh.

46	44 not 45

**Appendix 2. EMBASE search strategy**

1	pelvis tumor/

2	exp uterus cancer/

3	bladder tumor/

4	exp prostate tumor/

5	rectum tumor/

6	((pelvi* or cervi* or uter* or endometr* or bladder* or prostat* or rect* or anal or anus or genitourin* or genito-urin* or colorect* or gynaeco* or gyneco*) adj5 (cancer* or tumor* or tumour* or neoplas* or malignan* or carcinoma* or adenocarcinoma*)).mp.

7	1 or 2 or 3 or 4 or 5 or 6

8	exp radiotherapy/

9	rt.fs.

10	(radiotherap* or brachytherap* or radiat* or irradiat* or chemoradiotherap* or chemoradiat*).mp.

11	8 or 9 or 10

12	exp Probiotic/

13	exp Synbiotic/

14	exp Bacteria/

15	exp Prebiotic/

16	exp nutrition/

17	exp nutritional disorder/

18	exp polysaccharide/ or exp starch/

19	exp amino acid/

20	peptide/

21	exp antioxidant/

22	fish oil/

23	(nutrition* or nutrient* or malnutrition or diet* or food* or feed* or drink* or supplement* or formula* or vitamin* or mineral* or fat* or carbohydrate* or protein* or probiotic* or synbiotic* or bacteria* or prebiotic* or "amino acid*" or peptide*).mp.

24	(lactose or fibre or fiber or bran or psyllium or plantago ovata or ispaghula or polysaccharide* or polymeric* or peptide* or amino-acid* or glutamine* or fish oil* or arginine* or antioxidant* or anti-oxidant* or Metamucil or Inulin* or fructose-oligosaccharide* or fructo-oligosaccharide* or oligosaccharide* or starch or saccharide*).mp.

25	(enteral* or nasogastric* or gastrostomy of jejunostomy or oral* or tube* or sip*).mp.

26	12 or 13 or 14 or 15 or 16 or 17 or 18 or 19 or 20 or 21 or 22 or 23 or 24 or 25

27	7 and 11 and 26

28	crossover procedure/

29	double-blind procedure/

30	randomized controlled trial/

31	single-blind procedure/

32	random*.mp.

33	factorial*.mp.

34	(crossover* or cross over* or cross-over*).mp.

35	placebo*.mp.

36	(double* adj blind*).mp.

37	(singl* adj blind*).mp.

38	assign*.mp.

39	allocat*.mp.

40	volunteer*.mp.

41	controlled clinical trial/

42	cohort analysis/

43	prospective study/

44	retrospective study/

45	exp comparative study/

46	observational study/

47	case study/

48	(cohort* or prospective* or retrospective* or observational or comparative or (case* adj5 series)).mp.

49	28 or 29 or 30 or 31 or 32 or 33 or 34 or 35 or 36 or 37 or 38 or 39 or 40 or 41 or 42 or 43 or 44 or 45 or 46 or 47 or 48

50	27 and 49

51	(exp Animal/ or Nonhuman/ or exp animal Experiment/) not Human/

52	50 not 51

**Appendix 3. CENTRAL search strategy**

#1	MeSH descriptor: [Pelvic Neoplasms] this term only

#2	MeSH descriptor: [Uterine Neoplasms] explode all trees

#3	MeSH descriptor: [Urinary Bladder Neoplasms] this term only

#4	MeSH descriptor: [Prostatic Neoplasms] this term only

#5	MeSH descriptor: [Rectal Neoplasms] explode all trees

#6	((pelvi* or cervi* or uter* or endometr* or bladder* or prostat* or rect* or anal or anus or genitourin* or genito-urin* or colorect* or gynaeco* or gyneco*) near/5 (cancer* or tumor* or tumour* or neoplas* or malignan* or carcinoma* or adenocarcinoma*))

#7	#1 OR #2 OR #3 OR #4 OR #5 OR #6

#8	MeSH descriptor: [Radiotherapy] explode all trees

#9	MeSH descriptor: [] explode all trees and with qualifier(s): [radiotherapy - RT]

#10	radiotherap* or brachytherap* or radiat* or irradiat* or chemoradiotherap* or chemoradiat*

#11	#8 or #9 or #10

#12	MeSH descriptor: [Diet] explode all trees

#13	MeSH descriptor: [Feeding Behavior] this term only

#14	MeSH descriptor: [Food] explode all trees

#15	MeSH descriptor: [Nutrition Therapy] explode all trees

#16	MeSH descriptor: [Nutrition Disorders] explode all trees

#17	MeSH descriptor: [Feeding Methods] explode all trees

#18	MeSH descriptor: [Polysaccharides] explode all trees

#19	MeSH descriptor: [Starch] explode all trees

#20	MeSH descriptor: [Amino Acids] explode all trees

#21	MeSH descriptor: [Peptides] explode all trees

#22	MeSH descriptor: [Antioxidants] explode all trees

#23	MeSH descriptor: [Fish Oils] explode all trees

#24	nutrition* or nutrient* or malnutrition or diet* or food* or feed* or drink* or supplement* or formula* or vitamin* or mineral* or fat* or carbohydrate* or protein* or probiotic* or synbiotic* or bacteria* or prebiotic* or "amino acid*" or peptide*

#25	lactose or fibre or fiber or bran or psyllium or plantago ovata or ispaghula or polysaccharide* or polymeric* or peptide* or amino-acid* or glutamine* or fish oil* or arginine* or antioxidant* or anti-oxidant* or Metamucil or Inulin* or fructose-oligosaccharide* or fructo-oligosaccharide* or oligosaccharide* or starch or saccharide*

#26	enteral* or nasogastric* or gastrostomy or jejunostomy or oral* or tube* or sip*

#27	MeSH descriptor: [Probiotics] explode all trees

#28	MeSH descriptor: [Prebiotics] explode all trees

#29	MeSH descriptor: [Synbiotics] explode all trees

#30	MeSH descriptor: [Bacteria] explode all trees

#31	#12 OR #13 OR #14 OR #15 OR #16 OR #17 OR #18 OR #19 OR #20 OR #21 OR #22 OR #23 OR #24 OR #25 OR #26 OR #27 OR #28 OR #29 OR #30

#32	#7 AND #11 AND #31
